# A scoping review of fair machine learning techniques when using real-world data

**DOI:** 10.1101/2024.03.03.24303669

**Authors:** Yu Huang, Jingchuan Guo, Wei-Han Chen, Hsin-Yueh Lin, Huilin Tang, Fei Wang, Hua Xu, Jiang Bian

**Affiliations:** Department of Health Outcomes and Biomedical Informatics, University of Florida, Gainesville, FL, USA; Pharmaceutical Outcomes & Policy, University of Florida, Gainesville, FL, USA; Cornell University, USA; Yale University, USA

**Keywords:** Fair machine learning, bias mitigation, real-world data

## Abstract

**Objective:** The integration of artificial intelligence (AI) and machine learning (ML) in health care to aid clinical decisions is widespread. However, as AI and ML take important roles in health care, there are concerns about AI and ML associated fairness and bias. That is, an AI tool may have a disparate impact, with its benefits and drawbacks unevenly distributed across societal strata and subpopulations, potentially exacerbating existing health inequities. Thus, the objectives of this scoping review were to summarize existing literature and identify gaps in the topic of tackling algorithmic bias and optimizing fairness in AI/ML models using real-world data (RWD) in health care domains.

**Methods:** We conducted a thorough review of techniques for assessing and optimizing AI/ML model fairness in health care when using RWD in health care domains. The focus lies on appraising different quantification metrics for accessing fairness, publicly accessible datasets for ML fairness research, and bias mitigation approaches.

**Results:** We identified 11 papers that are focused on optimizing model fairness in health care applications. The current research on mitigating bias issues in RWD is limited, both in terms of disease variety and health care applications, as well as the accessibility of public datasets for ML fairness research. Existing studies often indicate positive outcomes when using pre-processing techniques to address algorithmic bias. There remain unresolved questions within the field that require further research, which includes pinpointing the root causes of bias in ML models, broadening fairness research in AI/ML with the use of RWD and exploring its implications in healthcare settings, and evaluating and addressing bias in multi-modal data.

**Conclusion:** This paper provides useful reference material and insights to researchers regarding AI/ML fairness in real-world health care data and reveals the gaps in the field. Fair AI/ML in health care is a burgeoning field that requires a heightened research focus to cover diverse applications and different types of RWD.

## Introduction

Recent advancements in artificial intelligence (AI) methods, especially machine learning (ML) algorithms, have led to successful integration into healthcare applications, demonstrating promising performance in various problems, such as risk prediction^1–3^, disease diagnosis^4–8^, and disease subphenotyping^9,10^. On the other hand, the rapid adoption of electronic health record (EHR) systems has made large collections of real-world data (RWD)^11,12^ available for research. There is an increasing interest in using these RWD, such as EHRs, administrative claims, and billing data, that are collected through routine care, to generate real-world evidence (RWE). Regulatory agencies such as the U.S. Food and Drug Administration (FDA) have started to issue guidance on how they consider RWD and RWE to support their regulatory decision-making^13^ and has indeed approved the new use of transplant drug based on observational RWD.^14^ The utilization of AI/ML-based methods, combined with readily available RWD, yields numerous benefits, e.g., more accurate disease and outcome prediction^15,16^ and efficient disease risk factors identification^17,18^, by effectively learning and analyzing patterns in a large collection of RWD^19^. However, despite the advantages of AI/ML techniques, there is a growing concern that such techniques may lead to unintended consequences, such as making biased decisions towards socioeconomically disadvantaged groups—leading to algorithmic fairness or bias issues.^20^

Algorithmic fairness is a relatively new research field in AI/ML but has rapidly gained increasing attention in recent years. A classic example of algorithmic bias comes from the Correctional Offender Management Profiling for Alternative Sanctions (COMPAS), a tool used by courts in the USA to identify the risk of a person recommitting another crime. The COMPAS has higher positive rates for African-American offenders than Caucasian offenders, often falsely predicting them to have a higher risk of recommitting a crime or recidivism.^21^ Similar examples have been found in the health care domain, such as the health care cost predictions are biased in the black and white groups^22^ and the presence of atherosclerotic disease being different across racial-ethnic groups^23^, the performance disparity across racial-ethnic and socioeconomic subgroups in ML models that predict heart failure outcomes^24^, and the ML-models producing unequal predictions in postpartum depression and postpartum mental health service utilization between White and African-American women^25^. The biased outputs of the ML models often stem from the inherent hidden biases in the data that these algorithms are trained on, reflecting the existing disparities in the society that generated these data^26^, and leading to exacerbating health equity issues, which negatively affects the utility, trustworthiness, and ethical use of ML models.

In recent years, the AI/ML research community has proposed a series of fairness assessments and subsequent mitigation techniques, which have been quickly adopted in solving various algorithmic bias issues in different applications. In the health and biomedical domain, there also exists some research investigating fairness issues when using AI/ML methods. In 2018, Gianfrancescogian et al^27^ discussed the potential bias in ML models using EHRs, and pointed out that the sources of bias in EHRs can be from missing data and patients missed by identification algorithms, sample size and potential underestimation, and misclassification or measurement error. There is a larger concern that biases or deficiencies in the EHR data used to train the ML algorithms and subsequently used in clinical decision support systems may contribute to disparities in health care.^27^ In 2020, Fletcher et al^28^ presented a few case studies and put forth a few important considerations on bias and fairness related to applying ML models in the context of global health, especially for low- and middle-income countries. In 2021, Mehrabi et al^26^ summarized how biases in data can skew what is learned by ML methods and comprehensively went over the definitions of fairness and bias that have been proposed in the literature. Moreover, they organized and established a taxonomy (pre-processing, in-processing, and post-processing) detailing the efforts undertaken to tackle or mitigate issues related to ML fairness across different domains. In 2022, Xu et al^20^ provided a scoping review about fair ML evaluation metrics in computational medicine, while Wan et al (2023)^29^ focused on the general modeling design for fair ML and went over the state-of-the-art in-processing techniques for improving ML fairness. Nevertheless, there has not been a comprehensive review that systematically summarized existing literature on not only ML fairness assessment but also the bias mitigation strategies when ML techniques are applied to RWD in the health care domain. Therefore, in this paper, we aimed to fill this important gap and provide an overview of the existing literature focused on assessing fairness, identifying potential sources of bias, and bias mitigating techniques in ML models using RWD.

## Methods

### Search strategy and selection criteria

The review process is divided into two rounds: the evaluation of existing relevant review articles and the assessment of individual studies. **Figure 1** summarizes the overall search and screening process. We first identified review articles to study state-of-the-art progress of fairness mitigation techniques, and then we collected and analyzed individual studies that focus on applying fair ML techniques in health care applications. In both phases, we adhered to the same methodology, which encompasses conducting a comprehensive literature search, reviewing abstracts and full texts, and extracting data from the selected articles. To analyze and study the latest cutting-edge fair ML techniques as presented in review articles, we collected peer-reviewed publications from mainstream computer science databases, including IEEE Xplore (n = 92), ACM Digital Library (n = 78), and Web of Science (n = 249), as state-of-the-art ML fairness assessment and bias mitigation strategies are mostly developed by computer scientists, using the search query: (“survey” OR “review” OR “overview”) AND (“unfairness” OR “fairness”) AND (“machine learning” OR “deep learning” OR “algorithm” OR “clustering” OR “artificial intelligence”) with filters limiting to studies published in the last decade. We then excluded the papers that (1) were not review articles, and (2) were not related to fairness in machine learning. All papers underwent a 2-person review process for inclusion in the manuscript.

**Figure 1.**
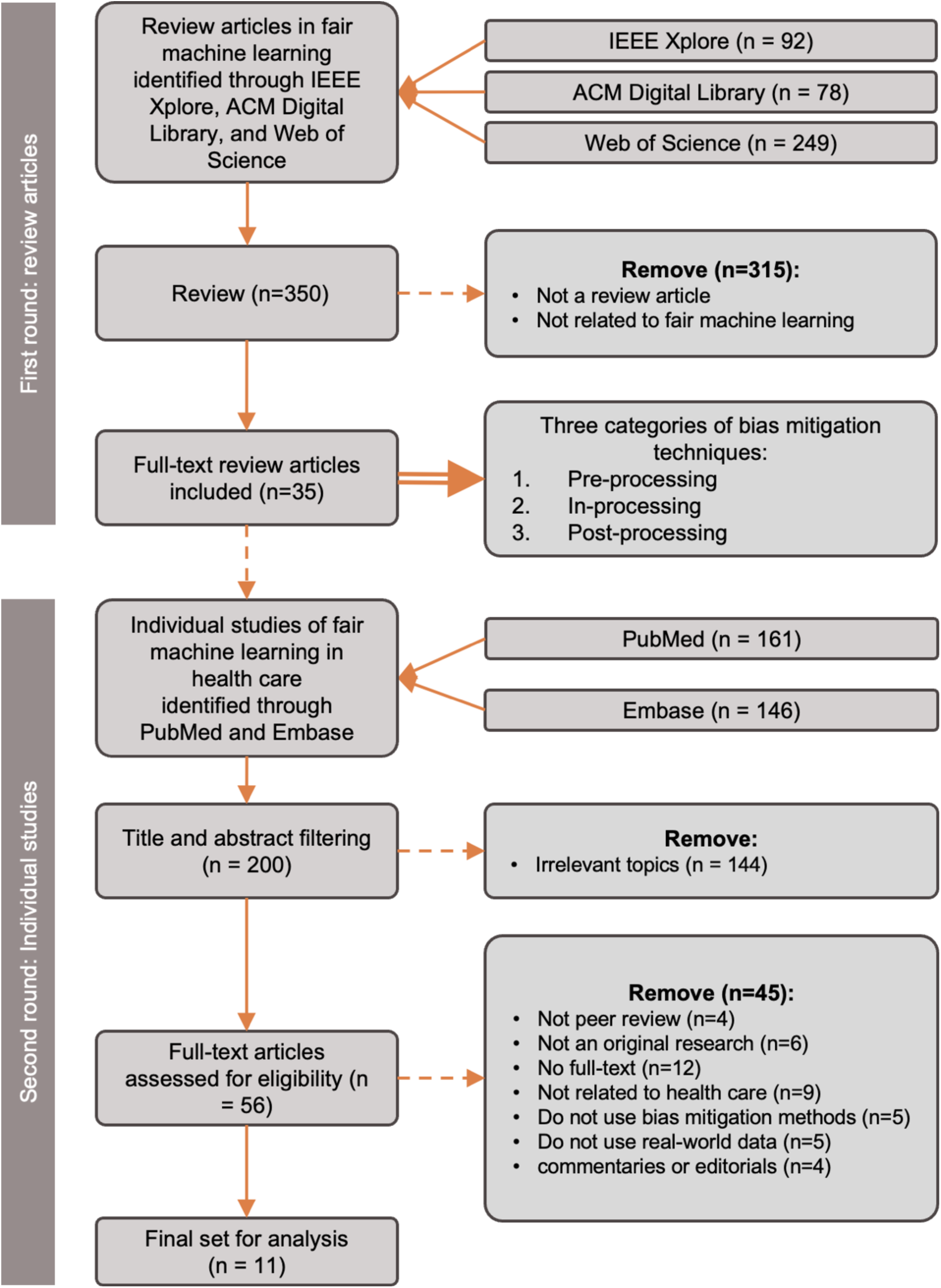
The overall search and screening process. MIMIC-III (Medical Information Mart for Intensive Care III), iSTAGING (Imaging-based coordinate SysTem for AGing and NeurodeGenerative diseases), GWTG-HF (Get With The Guidelines-Heart Failure), (NPD) National Pupil Database linked with the SLaM CAMHS (South London and Maudsley National Health Service Foundation Trust Child and Adolescent Mental Health Services) *Non-public dataset

In the second round, we extracted individual studies of fair ML in health care domain focusing on those using RWD, identified through PubMed (n=161) and Embase (n=146). Our initial inclusion criteria were studies that are (1) peer-review original research (excluding pre-prints), (2) having full-text available, (3) published within the last 10 years (encompassing years 2012–2022), (4) published in English, and (5) on the topic of mitigating bias issues in ML models using RWD. We excluded (1) articles that are not in English, (2) commentaries or editorials, (3) studies that did not use RWD, (4) studies that did not use bias mitigation methods, or (5) studies that are not in the health care domain. We followed the definition of RWD used by the FDA^13^ but focused on data generated from routine health care including EHRs, claims and billing data, and product and disease registries, but not including data collected in interventional, controlled, experimental clinical research settings. RWD also has other patient-generated health data, such as those generated in home-use care settings and data from mobile devices that can inform health status. We excluded studies using data that is generated from personal devices, such as smartphones and activity trackers, where different ML models are needed for these patient-generated health data.

## Results

As depicted in **Figure 1**, we identified 35 review articles discussing the topic of fair ML along with the fairness assessment and bias mitigation approaches. **Table 1** provides an overview of the array of reported algorithmic bias mitigation techniques. The search for individual studies yielded 11 distinct studies centered around the utilization of RWD for health care applications on the topic of fair ML.

**Table 1.** Overview of the existing bias mitigation approaches.

| Pre-processing | In-processing | Post-processing |
| --- | --- | --- |
| • Synthetic data generation<br>• Sensitive feature removal<br>• Sensitive feature replacement<br>• Reweighting<br>• Resampling<br>• Diverse data collection<br>• Adjusting label selection<br>• Disparate impact remover | • Regularization (e.g., Prejudice Remover)<br>• Constraint optimization<br>• Adversaria learning without generated data<br>• Adversarial learning with generated data | • Transformation<br>• Thresholding<br>• Constraint optimization<br>• Calibration<br>• Varying cut-point selection<br>• Reject option-based classification |

### Fairness assessment

An important first step in fair ML is to quantify the bias generated from various sources that affect the model. In this section, we summarized ten metrics that can be used to evaluate algorithmic fairness in the context of health care. The mathematical notations that are used in the formula of the metrics are listed in **Table 2**. We used a case study to illustrate these metrics, where the task was to build an ML model for predicting patient hospitalization risk using EHRs. The sensitive attribute was race i.e., Black (the protected group) versus White (the reference or privileged group), and the objective was to assess if the ML prediction model for hospitalization risk treated the two subgroups fairly.

**Table 2.** Notations and symbols.

| Symbol | Description |
| --- | --- |
| $G$ | Sensitive attribute |
| $Y$ | Actual outcome |
| $\hat{Y}$ | Predicted outcome |

***<u>Overall accuracy equality</u>*** (a.k.a. delta accuracy) is the evaluation of whether both the protected and privileged groups exhibit an equal level of prediction accuracy. Prediction accuracy, in this context, pertains to the likelihood of correctly assigning individuals from either a positive or negative class to their respective categories. The formula for overall accuracy equality in our case is shown below. ^30^

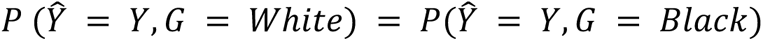

***<u>Equality of opportunity</u>*** (a.k.a. equal opportunity or false negative rate [FNR] balance or delta true positive rate) assesses whether both groups have equal FNR. The FNR refers to the probability of a subject belonging to the positive class being incorrectly predicted as negative. Notably, a model with equal FNRs will have an equal true positive rate (TPR) as well.^31^

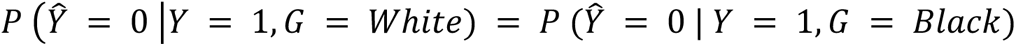

***<u>Predictive parity</u>*** is the concept where both the protected and privileged groups have an equal positive predictive value (PPV), which represents the probability of a subject, identified as positive, actually belonging to the positive class. Notably, a model with equal PPVs will have an equal false discovery rate (FDR) as well. ^31^

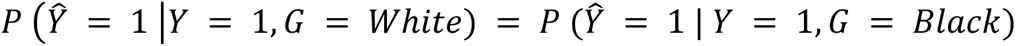

***<u>Predictive equality</u>***, also known as false positive rate (FPR) balance or delta false positive rate, refers to the condition where both the protected and privileged groups have an equal FPR. The FPR is the probability of a subject in a negative class being incorrectly predicted as positive. ^32,33^

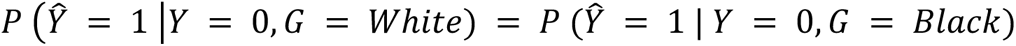

***<u>Statistical or demographic parity</u>*** is one of the earliest definitions of fairness, which is defined as each subgroup having an equal probability to be classified with a positive label. ^30,33^

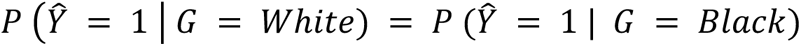

***<u>Disparate impact</u>*** examines the probability of subjects from different groups being classified with the positive label, which is similar to statistical/demographic parity. ^34^

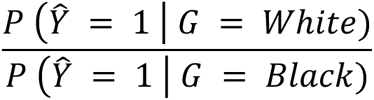

***<u>Equalized Odds,</u>*** also known as conditional procedure accuracy equality or disparate mistreatment, is a fairness metric that takes into account both TPR and FPR simultaneously. ^30^

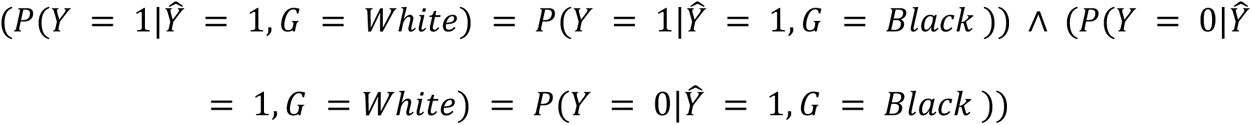

***<u>Intergroup standard deviation (IGSD)</u>*** is setup to understand the tradeoff between different metrics across multiple groups. Given a threshold *i*, metric *M*, and groups *A*, the IGSD*_Mi_* is defined as. ^35^

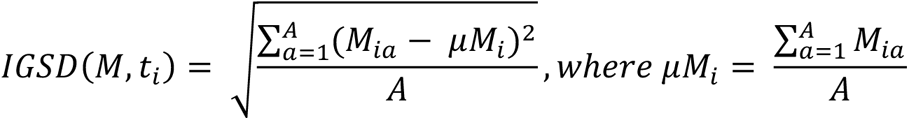

**<u>Conjunctive accuracy improvement (*CAI*_*α*_)</u>** is a combination of overall accuracy and accuracy gap (i.e., the difference in accuracy between subgroups) for measuring the success of mitigation algorithms. ^36^

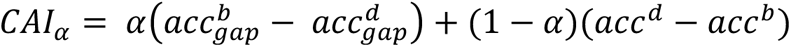

where *α* is the weight coefficient to trade-off the accuracy and fairness metrics, and *acc^b^* and 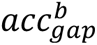 denote the accuracy and accuracy gap of the baseline model, and *acc^d^* and 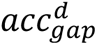 represent the accuracy and gap metrics of the debiased model, respectively. This metric can be generalized to other metrics (*CAUCI*_*α*_), using AUC as the performance metric.

**<u>Generalized entropy index (GEI)</u>** is to measure how unequally the outcomes of an algorithm benefit different individuals and subgroups. ^37^

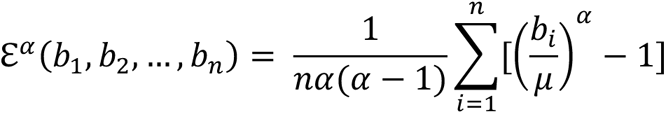

where *b_i_* is the benefit function of an individual *i* for presenting the predictive bias, and constant *α* ∉ {0,1} is a weight parameter.

### Algorithmic bias mitigation

The current review was focused on bias mitigation techniques on supervised ML models within the realm of health care. Readers who are interested in unsupervised learning algorithms can refer to this overview paper.^38^ As shown in **Figure 2**, the existing studies covered diverse disease domains and clinical applications, such as attention deficit hyperactivity disorder (ADHD)^39^, Alzheimer’s disease diagnosis^40^, cardiovascular disease prediction^41,42^, acute postoperative pain prediction^43^, survival analysis and mortality prediction^24,44^, postpartum depression (PPD) recognition^25^, diabetic retinopathy analysis^36^, and opioid misuse prediction ^45^.

**Figure 2.**
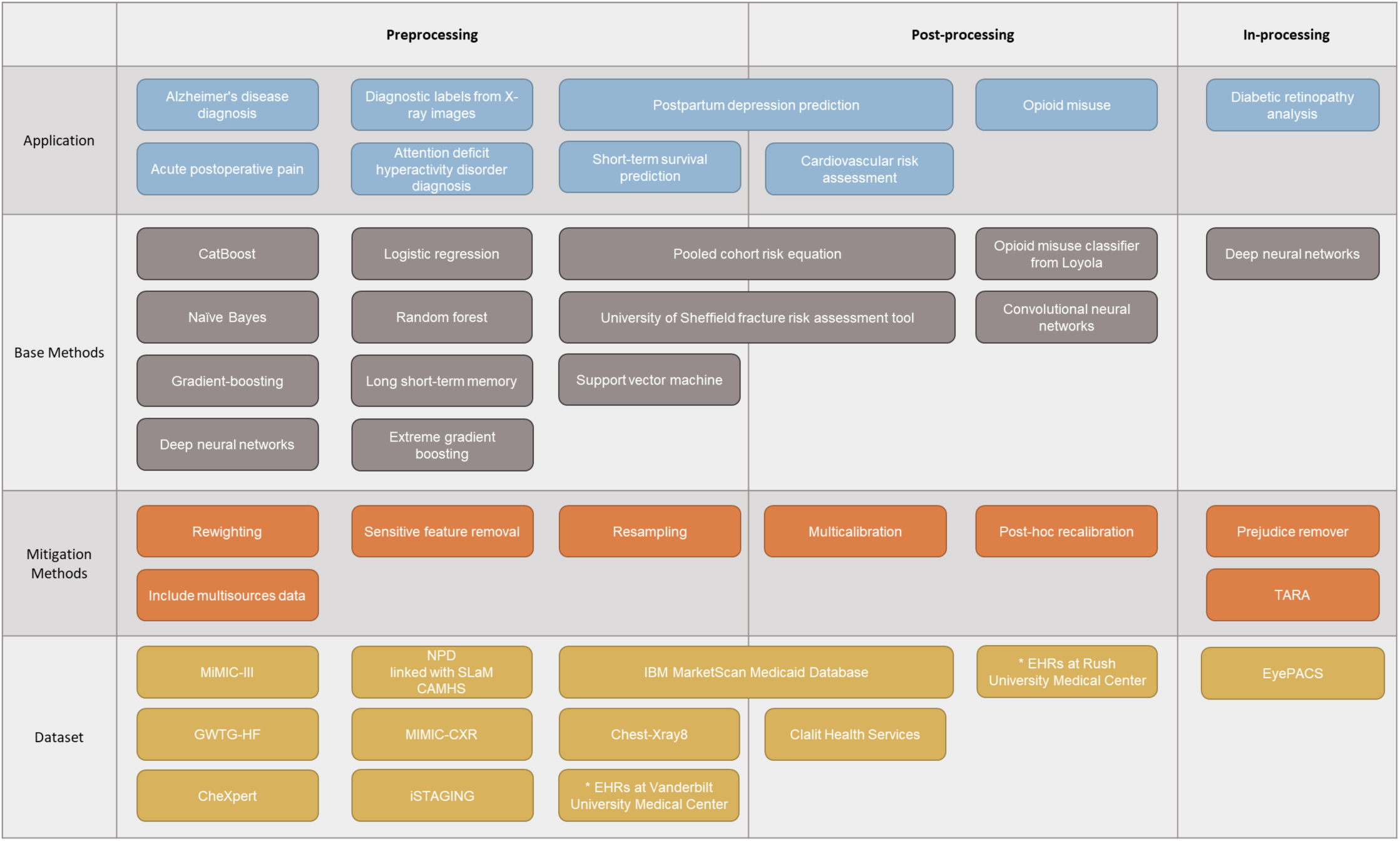
Fair machine learning in health care. MIMIC-III (Medical Information Mart for Intensive Care III), iSTAGING (Imaging-based coordinate SysTem for AGing and NeurodeGenerative diseases), GWTG-HF (Get With The Guidelines-Heart Failure), (NPD) National Pupil Database linked with the SLaM CAMHS (South London and Maudsley National Health Service Foundation Trust Child and Adolescent Mental Health Services) *Non-public dataset

Among these studies, eight conducted experiments using publicly accessible RWD datasets (ten in total), and **Figure 3** shows the breakdown of the data types. These datasets include the widely used Medical Information Mart for Intensive Care III (MIMIC III)^46^, Clalit Health Services (CHS) from Israel^47^, Get With The Guidelines-Heart Failure (GWTG-HF) registry dataset^48^, IBM MarketScan Medicaid Database^49^, EyePACS^50^, MIMIC-CXR^51^, CheXper^52^, Chest-Xray8^53^, Imaging-based coordinate SysTem for AGing and NeurodeGenerative diseases (iSTAGING) consortium^54^, and the National Pupil Database (NPD)^55^ linked with the South London and Maudsley National Health Service Foundation Trust Child and Adolescent Mental Health Services (SLaM CAMHS)^56,57^.

**Figure 3.**
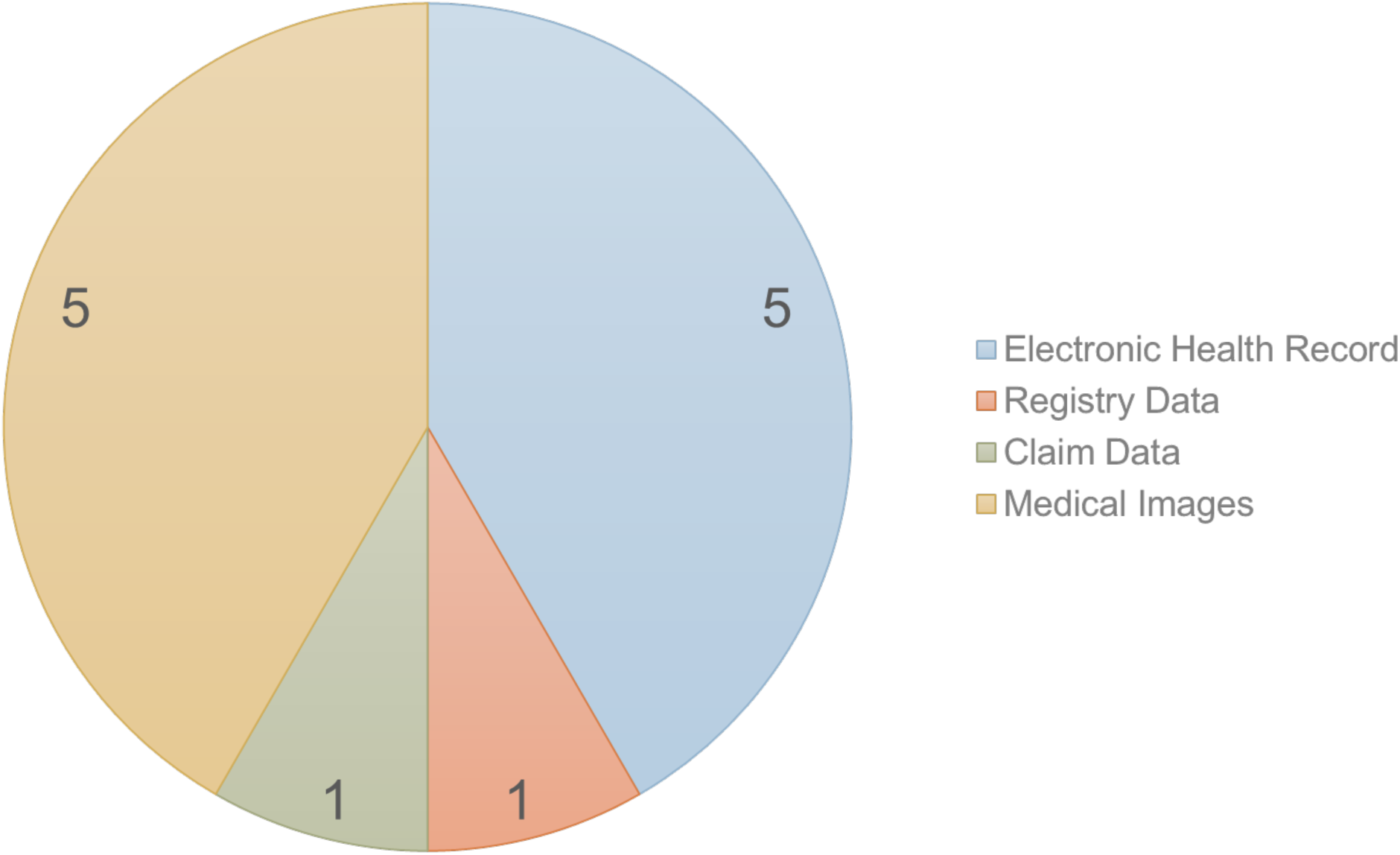
Distributions of data types of the real-world datasets used for fair ML research in health care.

The existing used datasets exhibit potential biases due to their specific geographical locations, contexts, and the populations they represent. For example, the datasets from Rush University Medical Center^45^ reflect the characteristics of local populations of Chicago, with White comprising 43.25% (n = 23,345), Black 32.50% (n=17,541), Hispanic/ Latinx 17.14% (n = 9252), and Other 7.11% (n = 3,836). The dataset from the University of Florida Health system^43^ covers the patients from Gainesville—a mid-size town with a mixture of young and older adults—and Jacksonville—a large city, characterized by a higher proportion of elderly individuals (average age = 60.72), a majority white population (79.96%) compared to non-whites (17.07%), a largely non-Hispanic group (93.35%) versus Hispanic (3.20%). Meanwhile, the dataset from Vanderbilt University Medical Center^42^ is limited to Black (13.7%) and White (86.3%). The NPD with the SLaM CAMHS dataset may be skewed towards the child and adolescent demographic in South London. Similarly, the CHS dataset from an Israeli health service could have biases reflecting Israeli demographics. The IBM MarketScan Medicaid Database, despite covering multiple states, is limited to low-income populations who are eligible for Medicaid. MIMIC III tends to represent critically ill patients in intensive care units and iSTAGING consortium is skewed toward elderly people.

Current approaches to bias mitigation strategies for ML ^26^ can be categorized into three groups (**Table 1**): Pre-processing, In-processing, and Post-processing. Based on our review, most studies (82%) used pre-processing methods to mitigate identified bias in clinical and biomedical applications.

#### Pre-processing

We identified nine papers that applied the pre-processing approaches for ML bias mitigation. Davoudi et al^43^ investigated the issue of prediction bias in CatBoost models designed to forecast acute postoperative pain. They employed a pre-processing technique known as “*reweighing*” to modify the weight of protected attributes, aiming to diminish the algorithmic bias in the prediction models. However, they found that reweighing the prediction models based on each protected attribute helped reduce the bias (for the predictive equality from 1.75 to less than 1.25) for some cases (e.g., lowest area deprivation index [ADI] tertile vs middle and highest tertiles), but it introduced bias in some other cases (English-speaking subgroup vs non-English speaking subgroup) where there was no bias (for the predictive equality from 0.84 to 0.29). Jeanselme et al^44^ demonstrated the impact of missing data in observational datasets on algorithmic fairness in subsequent stages. They conducted experiments using the MIMIC-III dataset to predict short-term survival within 7 days after an observation period. By comparing different imputation strategies (Multiple Imputation by Chained Equations [MICE] and group MICE), they revealed that prevalent imputation practices could negatively affect health equity, disproportionately affecting marginalized groups. Li et al^42^ investigated various predictive models, including Pooled Cohort Risk Equation (PCE), logistic regression (LR), random forest (RF), gradient-boosting trees, and long short-term memory (LSTM), intended for cardiovascular risk assessment based on extensive EHRs from Vanderbilt University Medical Center. They also showcased the potential of pre-processing methods such as eliminating sensitive attributes and resampling to mitigate the algorithmic bias originating from predictive models. Li et al ^24^ developed ML models to predict the probability of long-term hospitalization and in-hospital mortality for heart failure patients using admission data. They introduced a method to reduce models’ bias by integrating two sources of social determinants of health (SDoH), Social Deprivation Index (SDI)^58^ and ADI^59^, to build ML classifiers. Compared to the baseline (random forest [RF]) model, the model trained by using all SDoHs improves the demographic parity from 0.828 to 0.881 and the equalized odds ratio from 0.826 to 0.863. Wang et al. ^40^ used data from several magnetic resonance image (MRI) consortia for diagnosing Alzheimer’s disease, schizophrenia, and autism spectrum disorder. The authors evaluated the fairness of their deep neural network across subjects from different genders (female, male), ages (various cutoffs for different diseases), racial groups (various categories for different diseases), and clinical studies (various trials). They found that using multisource data (using demographics, clinical variables, genetic factors, and cognitive scores in addition to MRI features) improves the average AUC and achieves unbiased predictions (e.g., the difference of the equality of opportunity from 0.159 to 0.043 for Alzheimer’s disease diagnosis) across different subgroups of models for these three disorders.

Another work by Park et al^25^ tackled racial bias in predicting PPD and mental health service utilization using ML models trained on observational data. They explored various bias-mitigating techniques, including reweighing, prejudice remover, and removing race from the models. Reweighing was identified as a viable approach, enhancing fairness (e.g., for predicting PPD development, disparate impact improved from 0.31 to 0.79 and equality of opportunity improved from −0.19 to 0.02) without compromising accuracy (e.g., prediction accuracy from 0.73 to 0.72) across experimental conditions, unlike prejudice remover (prediction accuracy decreased from 0.73 to 0.68). Seyyed-Kalantari et al^60^ evaluated cutting-edge deep neural network models for predicting diagnostic labels from X-ray images collected through routine care. Their experiments suggested that employing multi-source datasets for training could counteract bias during data collection. Ter-Minassian et al^39^ investigated the prediction of Attention-Deficit Hyperactivity Disorder (ADHD) risk using ML and statistical methods with data from the NPD linked with SLaM CAMHS. To mitigate bias, they employed the pre-processing method of reweighing, which significantly improved disparate impact (from 0.15-0.60 to 1) without compromising the models’ ADHD classification performance.

#### Post-processing

A total of two papers employed the post-processing approaches for ML bias mitigation. Barda et al^41^ evaluated two baseline models, the Pooled Cohort Equations (PCE) and the University of Sheffield fracture risk assessment tool (FRAX), on the prediction of primary cardiovascular disease, and found that the post-processing algorithm (i.e., multicalibration^61^) improves model fairness (e.g., the difference between the mean observed and the mean predicted risk improved from 1.25 to 1.04 in the PCE and from 1.08 to 1.00 in the FRAX model) in subpopulations defined by 5 protected variables: ethnicity, sex, age group, socioeconomic status, and immigration status. Thompson^45^ assessed the fairness of a previously validated ML opioid misuse classifier based on the EHR data from Rush University Medical Center. Post-hoc recalibration methods were conducted to eliminate bias in FNR (from 0.32 to 0.24 in the black group) with minimal changes in other subgroup (from 0.17 to 0.21 in the white group) error metrics.

#### In-processing

In-processing techniques were less commonly used compared to pre-processing approaches in the existing health care literature. Two were identified, and only one paper shows the superiority of in-processing techniques. Paul et al^36^ proposed a novel debiasing approach that uses generative models and adversarial learning, called training and representation alteration (TARA), to improve algorithmic fairness. The proposed TARA approach was tested on several healthcare-related tasks, including evaluation of the state of mental health and diabetic retinopathy analysis, and the results demonstrate the superiority of fairness mitigation of the TARA compared with the competing baselines (e.g., the gap of accuracy reduced from 10.5 to 0.5 across different race groups on the diabetic retinopathy analysis task).

## Discussion

The existing state of research in fair ML utilizing RWD has primarily concentrated on diverse applications and disease domains as demonstration use cases. These studies have certainly illustrated the potential of fair ML in tackling a significant concern in AI-based clinical decision support — AI/ML producing biased predictions and thus exacerbating existing disparities. Our review identified several health care applications in fair ML research using RWD. The existing studies have undoubtedly highlighted the transformative potential of fair ML in different health care applications, but the advancement of this field requires a diverse array of data sources that encompass a wide range of health conditions. However, it is important to acknowledge that the range of applications discovered through our scoping review remains limited. It is crucial to broaden our endeavors to include a wider array of diseases and health-related concerns, thereby maximizing the impact of fair ML in minimizing healthcare inequalities and enhancing disease outcomes.

Our research identified ten fairness metrics currently employed in RWD studies. Most of these studies focused on group fairness metrics, such as predictive equality and equalized odds, while individual fairness measurement received less attention. As Kleinberg et al.^62^ noted, achieving different fairness metrics simultaneously is challenging, except in a few specific situations. This difficulty arises because these metrics focus on different aspects, and improving one metric often leads to a reduction in others. The choice of fairness metrics can influence the perceived fairness of clinical prediction models, highlighting the importance of context-based selection of metrics. Beyond the metrics themselves, it is crucial to consider whether biases in ML algorithms are solely technical issues, as the underlying biological and socioeconomic factors (e.g., social determinants of health) may be the root causes of the differences or disparities and contribute to the development of biases. This also leads to an interesting potential use of fairness metrics for assessing health disparities, where the underlying disparities were captured in the data and quantitatively reflected in the differences in the “fairness” metrics across, for example, racial and ethinical groups and socioeconomically vulnerable populations. Therefore, collaboration with domain and legal experts is important to comprehensively understand the specific context of problems, evaluate the ethical considerations of using ML techniques, and select appropriate fairness metrics.

Our review identified n=9, 2, and 2 studies that employed pre-processing, in-processing, and post-processing techniques for ML bias mitigation. While pre-processing techniques have been widely used, there is a lack of evidence demonstrating their superiority over post-processing or in-processing alternatives. Comparative evaluations of the three different classes of ML mitigation approaches are an important topic but yet to be studied. To advance our understanding of the most effective strategies in addressing algorithmic biases within healthcare domains, there exists a need for comprehensive studies that systematically assess the strengths and weaknesses of pre-processing, post-processing, and in-processing approaches. By comprehensively evaluating the trade-offs and performance nuances of these techniques, researchers can shed light on the most suitable interventions for specific contexts and bolster the foundation for a fairer and more robust ML landscape within health care.

While existing studies have made notable strides in mitigating algorithmic bias within single-modality data like structured EHRs and medical imaging data, the emergence of multi-modality data in health care research presents both opportunities and challenges. The essential task of untangling and mitigating bias across different modalities and understanding the contributions of each modality to potential biases becomes increasingly complex. Furthermore, the underexplored terrain of unstructured data, including clinical narratives and textual data, holds immense potential for comprehensively analyzing patient health status. Unfortunately, the current landscape lacks sufficient methods or studies that effectively evaluate and mitigate algorithmic bias within these advanced models, especially when dealing with complex language models like Large Language Models (LLMs), in the context of clinical notes and other unstructured data. Advancing fair ML into the realm of multi-modality and unstructured data is an important research direction for promoting trustworthy AI in supporting the quality of health care.

Understanding model interpretation plays an important role in comprehending how a model works and in identifying the causes of the biases. While this understanding may not directly eliminate biases, it can help reduce biases in the decision-making process on how to use the models. Recently, explainable AI (XAI) has gained significance in bridging the gap between complex AI models and human understanding, particularly in healthcare^63–65^. For instance, Shapley Additive exPlanations (SHAP)^66^ is increasingly popular as a straightforward method for elucidating the contribution of individual factors to a predictive model. Alternatively, the attention mechanism effectively highlights the importance of each feature in deep learning models, such as those for EHRs^67^ and medical imaging^68^. Enhanced comprehension of model functionalities can better inform clinicians who use these models, leading to more equitable decision-making.

Applying bias mitigation methods to improve fairness in ML within the realm of clinical and biomedical research using RWD represents a transformative shift towards more ethical, precise, and patient-centered health care practices. The development and innovation of approaches and techniques to improve ML fairness and building trustworthy AI are critically needed across multifaceted aspects. First, fair ML can ensure clinical decision support are unbiased across diverse patient populations and social strata. Additionally, fair ML can not only improve the equity of healthcare processes and outcomes but also foster trust among healthcare providers, patients, and their families.

In addition to mitigating biases in AI/ML algorithms, it is necessary to pay attention to data collection, especially issues related to governance that affect how data can be collected, shared, and used, in healthcare systems, ensuring high-quality data to avoid biased data generation and collection. Mehrabi et al. ^26^ highlighted that the existing biases in AI systems can be divided into three categories (i.e., data, algorithms, and user interactions), and they interact in a feedback loop, influencing each other. Mitigating the biases in both models and data aspects has high potential to positively improve this feedback loop. Another issue is the lack of easy access to nationwide RWD databases with population diversity for building fair algorithms. Most health data used in model development comes from individual medical centers, which may not represent the population of the country, potentially leading to potential biases in the resulting models. Nevertheless, issues with generalizability stemming from the population representativeness of study samples are not unique to the development of AI/ML models; for example, they are widely discussed in the context of clinical trials.^69^ For developing AI/ML models, nationwide or even international data-sharing initiatives are essential to address these biases. Adopting a more liberal approach to promote data sharing across states or nations could help reduce these data biases, but it involves a trade-off between data privacy and data access. National and international research networks with large collections of RWD, such as the National Patient-Centered Clinical Research Network (PCORnet) funded by the Patient-Centered Outcomes Research Institute (PCORI), covering > 80 million patient records nationally,^70^ and the “open-source” Observational Health Data Sciences and Informatics (ODSRI) network, encompassing over 500 million patient records,^71^ combined with novel approaches like federated and distributed learning,^72^ would enhance access to representative data sample to mitigate such biases in AI/ML.

Looking ahead, there are several promising future research directions in addressing AI/ML fairness in clinical research using RWD. First, it is critical to develop a research framework and associated methods and tools that can reveal the underlying potential causes of bias in an ML algorithm. Moreover, existing research on fair ML is predominantly limited to regular ML algorithms and a relatively narrow range of healthcare applications. There is potential for expanding fair ML research into more advanced AI/ML algorithms (e.g., AutoML, deep learning) and broader healthcare data domains and applications (e.g., electrocardiogram waveform data). Additionally, given the increasing prominence of multimodality data in healthcare research, there is a pressing need for innovative techniques that can account for biases arising from interactions among different modalities.^73^ Similarly, recognizing the significance of unstructured data, such as clinical narratives, highlights the need for innovative fair ML methods specifically designed for these complex textual resources.

Our study had two major limitations. Firstly, we excluded studies and theses not written in English. Secondly, we only reviewed articles on the topic of fair ML using RWD in healthcare domains. Further investigation into how bias in RWD and analytic methods affects study results should be systematically conducted.

## Conclusion

Fair ML can offer equitable and impactful clinical decision-support tools to improve the quality of care and health outcomes. Our scoping review provides useful reference material and insights for researchers regarding AI/ML fairness in real-world healthcare data, revealing the gaps in the field. Fair AI/ML in healthcare is a burgeoning field that requires heightened research focus to cover diverse applications and various types of RWD.

## Data Availability

All data produced in the present work are contained in the manuscript

## Notes

### Competing Interest Statement

The authors have declared no competing interest.

## Reference

1. Pudjihartono N, Fadason T, Kempa-Liehr AW, O’Sullivan JM. A Review of Feature Selection Methods for Machine Learning-Based Disease Risk Prediction. Front Bioinform. 2022;2:927312.

2. Suri JS, Bhagawati M, Paul S, et al. Understanding the bias in machine learning systems for cardiovascular disease risk assessment: The first of its kind review. Comput Biol Med. 2022;142:105204.

3. Li Q, Yang X, Xu J, et al. Early prediction of Alzheimer’s disease and related dementias using real-world electronic health records. Alzheimers Dement. Published online February 23, 2023. doi:10.1002/alz.12967

4. Kononenko I. Machine learning for medical diagnosis: history, state of the art and perspective. Artif Intell Med. 2001;23(1):89–109.

5. Bakator M, Radosav D. Deep Learning and Medical Diagnosis: A Review of Literature. Multimodal Technologies and Interaction. 2018;2(3):47.

6. Petrova-Antonova D, Spasov I, Krasteva I, Manova I, Ilieva S. A Digital Twin Platform for Diagnostics and Rehabilitation of Multiple Sclerosis. In: Computational Science and Its Applications – ICCSA 2020. Springer International Publishing; 2020:503–518.

7. Battineni G, Sagaro GG, Chinatalapudi N, Amenta F. Applications of Machine Learning Predictive Models in the Chronic Disease Diagnosis. J Pers Med. 2020;10(2). doi:10.3390/jpm10020021

8. Ahsan MM, Luna SA, Siddique Z. Machine-Learning-Based Disease Diagnosis: A Comprehensive Review. Healthcare (Basel). 2022;10(3). doi:10.3390/healthcare10030541

9. Zhang H, Zang C, Xu Z, et al. Data-driven identification of post-acute SARS-CoV-2 infection subphenotypes. Nat Med. Published online December 1, 2022:1–10.

10. Xu J, Bian J, Fishe JN. Pediatric and Adult Asthma Clinical Phenotypes: A Real World, Big Data Study Based on Acute Exacerbations. J Asthma Allergy. Published online August 29, 2022:1–11.

11. Sherman RE, Anderson SA, Dal Pan GJ, et al. Real-world evidence - what is it and what can it tell us? N Engl J Med. 2016;375(23):2293–2297.

12. Concato J, Corrigan-Curay J. Real-world evidence - where are we now? N Engl J Med. 2022;386(18):1680–1682.

13. Center for Drug Evaluation, Research. Considerations for the use of real-world data and real-world evidence to support regulatory decision-making for drug and biological products. U.S. Food and Drug Administration. Published August 30, 2023. Accessed September 16, 2023. https://www.fda.gov/regulatory-information/search-fda-guidance-documents/considerations-use-real-world-data-and-real-world-evidence-support-regulatory-decision-making-drug

14. Center for Drug Evaluation, Research. FDA approves new use of transplant drug based on real-world evidence. U.S. Food and Drug Administration. Published September 30, 2021. Accessed January 24, 2023. https://www.fda.gov/drugs/news-events-human-drugs/fda-approves-new-use-transplant-drug-based-real-world-evidence

15. Shamout F, Zhu T, Clifton DA. Machine Learning for Clinical Outcome Prediction. IEEE Rev Biomed Eng. 2021;14:116–126.

16. Xie S, Yu Z, Lv Z. Multi-Disease Prediction Based on Deep Learning: A Survey. CMES-Computer Modeling in Engineering & Sciences. 2021;128(2). https://cdn.techscience.cn/uploads/attached/file/20210722/20210722075216_41143.pdf

17. Perveen S, Shahbaz M, Keshavjee K, Guergachi A. A Systematic Machine Learning Based Approach for the Diagnosis of Non-Alcoholic Fatty Liver Disease Risk and Progression. Sci Rep. 2018;8(1):2112.

18. Chatterjee A, Gerdes MW, Martinez SG. Identification of Risk Factors Associated with Obesity and Overweight—A Machine Learning Overview. Sensors. 2020;20(9):2734.

19. Office of the Commissioner. Real-World Evidence. U.S. Food and Drug Administration. Accessed May 19, 2023. https://www.fda.gov/science-research/science-and-research-special-topics/real-world-evidence

20. Xu J, Xiao Y, Wang WH, et al. Algorithmic fairness in computational medicine. eBioMedicine. 2022;84. doi:10.1016/j.ebiom.2022.104250

21. Angwin J, Larson J, Kirchner L, Mattu S. Machine bias. ProPublica. Published May 23, 2016. Accessed June 13, 2023. https://www.propublica.org/article/machine-bias-risk-assessments-in-criminal-sentencing

22. Obermeyer Z, Powers B, Vogeli C, Mullainathan S. Dissecting racial bias in an algorithm used to manage the health of populations. Science. 2019;366(6464):447–453.

23. Gijsberts CM, Groenewegen KA, Hoefer IE, et al. Race/Ethnic Differences in the Associations of the Framingham Risk Factors with Carotid IMT and Cardiovascular Events. PLoS One. 2015;10(7):e0132321.

24. Li Y, Wang H, Luo Y. Improving Fairness in the Prediction of Heart Failure Length of Stay and Mortality by Integrating Social Determinants of Health. Circ Heart Fail. 2022;15(11):e009473.

25. Park Y, Hu J, Singh M, et al. Comparison of Methods to Reduce Bias From Clinical Prediction Models of Postpartum Depression. JAMA Netw Open. 2021;4(4):e213909.

26. Mehrabi N, Morstatter F, Saxena N, Lerman K, Galstyan A. A Survey on Bias and Fairness in Machine Learning. ACM Comput Surv. 2021;54(6):1–35.

27. Gianfrancesco MA, Tamang S, Yazdany J, Schmajuk G. Potential Biases in Machine Learning Algorithms Using Electronic Health Record Data. JAMA Intern Med. 2018;178(11):1544–1547.

28. Fletcher RR, Nakeshimana A, Olubeko O. Addressing Fairness, Bias, and Appropriate Use of Artificial Intelligence and Machine Learning in Global Health. Front Artif Intell. 2020;3:561802.

29. Wan M, Zha D, Liu N, Zou N. In-Processing Modeling Techniques for Machine Learning Fairness: A Survey. ACM Trans Knowl Discov Data. 2023;17(3):1–27.

30. Berk R, Heidari H, Jabbari S, Kearns M, Roth A. Fairness in Criminal Justice Risk Assessments: The State of the Art. Sociol Methods Res. 2021;50(1):3–44.

31. Verma S, Rubin J. Fairness definitions explained. In: Proceedings of the International Workshop on Software Fairness. FairWare ‘18. Association for Computing Machinery; 2018:1–7.

32. Chouldechova A. Fair prediction with disparate impact: A study of bias in recidivism prediction instruments. Big Data. 2017;5(2):153–163.

33. Corbett-Davies S, Pierson E, Feller A, Goel S, Huq A. Algorithmic Decision Making and the Cost of Fairness. In: Proceedings of the 23rd ACM SIGKDD International Conference on Knowledge Discovery and Data Mining. KDD ‘17. Association for Computing Machinery; 2017:797–806.

34. Feldman M, Friedler SA, Moeller J, Scheidegger C, Venkatasubramanian S. Certifying and Removing Disparate Impact. In: Proceedings of the 21th ACM SIGKDD International Conference on Knowledge Discovery and Data Mining. KDD ‘15. Association for Computing Machinery; 2015:259–268.

35. Foryciarz A, Pfohl SR, Patel B, Shah N. Evaluating algorithmic fairness in the presence of clinical guidelines: the case of atherosclerotic cardiovascular disease risk estimation. BMJ Health Care Inform. 2022;29(1):e100460.

36. Paul W, Hadzic A, Joshi N, Alajaji F, Burlina P. TARA: Training and Representation Alteration for AI Fairness and Domain Generalization. Neural Comput. 2022;34(3):716–753.

37. Speicher T, Heidari H, Grgic-Hlaca N, et al. A Unified Approach to Quantifying Algorithmic Unfairness: Measuring Individual &Group Unfairness via Inequality Indices. In: Proceedings of the 24th ACM SIGKDD International Conference on Knowledge Discovery & Data Mining. KDD ‘18. Association for Computing Machinery; 2018:2239–2248.

38. Chhabra A, Masalkovaite K, Mohapatra P. An overview of fairness in clustering. IEEE Access. 2021;9:130698–130720.

39. Ter-Minassian L, Viani N, Wickersham A, et al. Assessing machine learning for fair prediction of ADHD in school pupils using a retrospective cohort study of linked education and healthcare data. BMJ Open. 2022;12(12):e058058.

40. Wang R, Chaudhari P, Davatzikos C. Bias in machine learning models can be significantly mitigated by careful training: Evidence from neuroimaging studies. Proceedings of the National Academy of Sciences. 2023;120(6):e2211613120.

41. Barda N, Yona G, Rothblum GN, et al. Addressing bias in prediction models by improving subpopulation calibration. J Am Med Inform Assoc. 2021;28(3):549–558.

42. Li F, Wu P, Ong HH, Peterson JF, Wei WQ, Zhao J. Evaluating and mitigating bias in machine learning models for cardiovascular disease prediction. J Biomed Inform. 2023;138:104294.

43. Davoudi A, Sajdeya R, Ison R, et al. Fairness in the prediction of acute postoperative pain using machine learning models. Front Digit Health. 2022;4:970281.

44. Jeanselme V, De-Arteaga M, Zhang Z, Barrett J, Tom B. Imputation Strategies Under Clinical Presence: Impact on Algorithmic Fairness. In: Parziale A, Agrawal M, Joshi S, et al., eds. Proceedings of the 2nd Machine Learning for Health Symposium. Vol 193. Proceedings of Machine Learning Research. PMLR; 2022:12–34.

45. Thompson HM, Sharma B, Bhalla S, et al. Bias and fairness assessment of a natural language processing opioid misuse classifier: detection and mitigation of electronic health record data disadvantages across racial subgroups. J Am Med Inform Assoc. 2021;28(11):2393–2403.

46. Johnson AEW, Pollard TJ, Shen L, et al. MIMIC-III, a freely accessible critical care database. Sci Data. 2016;3:160035.

47. Clalit Health Services. Published 2023. http://clalitresearch.org/about-us/

48. Smaha LA, American Heart Association. The American Heart Association Get With The Guidelines program. Am Heart J. 2004;148(5 Suppl):S46–8.

49. Hansen L. IBM MarketScan Research Databases for life sciences researchers. IBM Watson Health.

50. Cuadros J, Bresnick G. EyePACS: an adaptable telemedicine system for diabetic retinopathy screening. J Diabetes Sci Technol. 2009;3(3):509–516.

51. Johnson A, Pollard T, Mark R, Berkowitz S, Horng S. MIMIC-CXR Database (version 2.0. 0). PhysioNet. Published online 2019.

52. Irvin J, Rajpurkar P, Ko M, et al. CheXpert: A Large Chest Radiograph Dataset with Uncertainty Labels and Expert Comparison. AAAI. 2019;33(01):590–597.

53. Wang X, Peng Y, Lu L, Lu Z, Bagheri M, Summers RM. ChestX-Ray8: Hospital-scale chest X-ray database and benchmarks on weakly-supervised classification and localization of common thorax diseases. In: 2017 IEEE Conference on Computer Vision and Pattern Recognition (CVPR). IEEE; 2017:2097–2106.

54. Habes M, Pomponio R, Shou H, et al. The Brain Chart of Aging: Machine-learning analytics reveals links between brain aging, white matter disease, amyloid burden, and cognition in the iSTAGING consortium of 10,216 harmonized MR scans. Alzheimers Dement. 2021;17(1):89–102.

55. Jay MA, McGrath-Lone L, Gilbert R. Data Resource: the National Pupil Database (NPD). Int J Popul Data Sci. 2019;4(1):1101.

56. Downs J, Gilbert R, Hayes RD, Hotopf M, Ford T. Linking health and education data to plan and evaluate services for children. Arch Dis Child. 2017;102(7):599–602.

57. Downs JM, Ford T, Stewart R, et al. An approach to linking education, social care and electronic health records for children and young people in South London: a linkage study of child and adolescent mental health service data. BMJ Open. 2019;9(1):e024355.

58. Butler DC, Petterson S, Phillips RL, Bazemore AW. Measures of social deprivation that predict health care access and need within a rational area of primary care service delivery. Health Serv Res. 2013;48(2 Pt 1):539–559.

59. Kind AJH, Buckingham WR. Making Neighborhood-Disadvantage Metrics Accessible - The Neighborhood Atlas. N Engl J Med. 2018;378(26):2456–2458.

60. Seyyed-Kalantari L, Liu G, McDermott M, Chen IY, Ghassemi M. CheXclusion: Fairness gaps in deep chest X-ray classifiers. Pac Symp Biocomput. 2021;26:232–243.

61. Hebert-Johnson U, Kim M, Reingold O, Rothblum G. Multicalibration: Calibration for the (Computationally-Identifiable) Masses. In: Dy J, Krause A, eds. Proceedings of the 35th International Conference on Machine Learning. Vol 80. Proceedings of Machine Learning Research. PMLR; 10--15 Jul 2018:1939–1948.

62. Kleinberg J, Mullainathan S, Raghavan M. Inherent trade-offs in the fair determination of risk scores. arXiv [csLG]. Published online September 19, 2016. http://arxiv.org/abs/1609.05807

63. Saraswat D, Bhattacharya P, Verma A, et al. Explainable AI for Healthcare 5.0: Opportunities and Challenges. IEEE Access. 2022;10:84486–84517.

64. Loh HW, Ooi CP, Seoni S, Barua PD, Molinari F, Acharya UR. Application of explainable artificial intelligence for healthcare: A systematic review of the last decade (2011–2022). Comput Methods Programs Biomed. 2022;226:107161.

65. Payrovnaziri SN, Chen Z, Rengifo-Moreno P, et al. Explainable artificial intelligence models using real-world electronic health record data: a systematic scoping review. J Am Med Inform Assoc. 2020;27(7):1173–1185.

66. Lundberg S, Lee SI. A unified approach to interpreting model predictions. arXiv [csAI]. Published online May 22, 2017. Accessed January 15, 2023. https://proceedings.neurips.cc/paper/2017/hash/8a20a8621978632d76c43dfd28b67767-Abstract.html

67. Meng Y, Speier W, Ong M, Arnold CW. HCET: Hierarchical Clinical Embedding With Topic Modeling on Electronic Health Records for Predicting Future Depression. IEEE J Biomed Health Inform. 2021;25(4):1265–1272.

68. Selvaraju RR, Cogswell M, Das A, Vedantam R, Parikh D, Batra D. Grad-CAM: Visual Explanations from Deep Networks via Gradient-Based Localization. In: 2017 IEEE International Conference on Computer Vision (ICCV). IEEE; 2017:618–626.

69. He Z, Tang X, Yang X, et al. Clinical trial generalizability assessment in the big data era: A review. Clin Transl Sci. 2020;13(4):675–684.

70. Forrest CB, McTigue KM, Hernandez AF, et al. PCORnet® 2020: current state, accomplishments, and future directions. J Clin Epidemiol. 2021;129:60–67.

71. OHDSI. OHDSI – observational health data sciences and informatics. Published 2024. Accessed January 17, 2024. https://www.ohdsi.org/

72. Xu J, Glicksberg BS, Su C, Walker P, Bian J, Wang F. Federated learning for healthcare informatics. J Healthc Inform Res. 2020;5(1):1–19.

73. Booth BM, Hickman L, Subburaj SK, Tay L, Woo SE, D’Mello SK. Bias and Fairness in Multimodal Machine Learning: A Case Study of Automated Video Interviews. In: Proceedings of the 2021 International Conference on Multimodal Interaction. ICMI ‘21. Association for Computing Machinery; 2021:268–277.

